# The complexity of anticoagulant prescribing in advanced kidney disease: a case vignette study

**DOI:** 10.64898/2026.06.25.26356635

**Authors:** Kathrine Parker, Sandip Mitra, Jecko Thachil, Penny Lewis

## Abstract

**Background:** Anticoagulants are high-risk medications that require careful consideration of the risks and benefits. In the setting of advanced kidney disease decisions around prescribing anticoagulants is problematic. This relates to a lack of good-quality evidence and the altered renal clearance and protein binding in kidney disease. This leads to prescribing variation and results in patients experiencing different standards of care. The aim of this study was to explore the factors influencing anticoagulant prescribing decisions among nephrology and haematology healthcare professionals in commonly encountered scenarios, in order to better understand variation in prescribing practice.

**Methods:** Three case-vignettes were presented to 15 haematology and nephrology professionals as part of a semi-structured interview. Participants were purposively selected based on variable responses to a UK anticoagulant prescribing practice survey. An inductive thematic analysis was undertaken as described by Braun and Clark with an iterative review process to identify final themes.

**Results:** There were five main themes that arose from the analysis; evidence, patient factors, knowledge and experience, team influence and systems and they were present across all the three vignettes. The prescribing of anticoagulation in advanced kidney disease is strongly influenced by the knowledge and expertise gleaned by prescribers’ previous experiences and the incorporation of patient factors. Prescribers commonly seek opinion from senior staff and other members of the multiprofessional team to support their decision making. Decisions are also influenced by organisational factors that are beyond their individual control.

**Conclusion:** Development of guidelines, delivery of education and good-quality evidence is needed to improve prescribing variation and ultimately the quality of care provided to patients with advanced kidney disease.

## Introduction

Anticoagulants, including low molecular weight heparins (LMWH), vitamin K antagonists (VKA) and direct oral anticoagulants (DOACs) are used in the treatment of acute venous thromboembolism (VTE), atrial fibrillation to reduce the risk of stroke and for VTE thromboprophylaxis. Anticoagulants are considered high-risk medications requiring careful consideration of the risks and benefits of treatment with potential for serious bleeding events and thrombotic complications if not appropriately dosed(1). The prescribing of any medication can be complex and requires healthcare professionals to integrate the existing evidence base, their clinical expertise and individual patient circumstances and values to make a shared decision(2, 3).

In the case of anticoagulant prescribing in patients with advanced kidney disease, the decision to prescribe is especially problematic. This is due to multiple factors including a lack of, or poor quality, evidence: people with kidney disease are often excluded from randomised controlled studies (RCTs)(4). There is an absence of high-quality studies investigating anticoagulants in those with nephrotic syndrome(5), limiting the applicability of existing evidence. Furthermore, altered renal clearance and protein binding in advanced kidney disease makes their use more challenging. People with advanced kidney disease have a complex pathophysiology placing them at higher risk of thrombotic and bleeding events and balancing these risks can be challenging(6).

The existing evidence gap leads to a variation in prescribing, as reported in our previous survey of anticoagulant prescribing of UK haematology and nephrology professionals, in which prescribing practice varied not only between different hospitals but also within the same hospital(7). Prescribers find this variation difficult to navigate, particularly when many prescribers would rely on guidelines to support their prescribing decisions(8). It also results in patients experiencing different standards of care depending on where and by whom they are treated – this can lead to the reduced safety of anticoagulant use(9). Such variation can be confusing for patients who have reported a lack of clarity with regards to their anticoagulant plan(10) .

The underlying rationale for variability in anticoagulant prescribing decisions among prescribers are unclear. Yet this understanding could support the future development of recommendations or best-practice guidelines to standardise decision making in anticoagulation prescribing in advanced kidney disease and support clinicians and patients. This need was previously highlighted by a Canadian study that concluded that a guideline would be of great value to both nephrology professionals and patients(8).

Therefore, the aim of this study was to explore the factors influencing anticoagulant prescribing decisions among nephrology and haematology healthcare professionals in commonly encountered scenarios, in order to better understand variation in prescribing practice.

## Methods

### Study design

Case vignettes are short cases presented to individuals with knowledge of a topic to share their perceptions on clinical dilemmas they face (11). Case vignettes enable an in-depth exploration of health professionals’ decision making as participants’ can verbalise their decision-making process while working through the case, an approach called think-out loud methodology(12). This approach increases the quality of data as it does not rely on participant recall and it avoids generalisation(12). In this study, participants were asked to verbalise their decision-making process while working through clinical vignettes based on anticoagulation use in advanced CKD (13). A list of pre-formed questions were developed and asked by the interviewer (KP) following the vignettes to identify the reasons for their prescribing decisions. This included why a particular anticoagulant and dose was selected, if this information had not already been provided during the think-aloud process. The questions can be found in supplementary appendix 1.

Three case vignettes were designed by a renal pharmacist (KP) based on areas of prescribing variation from our previously published UK prescribing practice survey(7). These included 1) treatment of VTE in a patient with CKD stage 4 and systemic lupus erythematosus 2) the use of anticoagulation in a dialysis patient with AF who had developed calciphylaxis on warfarin and 3) anticoagulation use in a patient with acute nephrotic syndrome, supplementary appendix 1. The vignettes were reviewed by a consultant nephrologist (SM) and consultant haematologist (JT). A nephrology resident doctor (JH) piloted the vignettes and provided feedback which included providing a diagnosis for the nephrotic syndrome as that may affect anticoagulation decision making.

### Participant recruitment

Nephrology and haematology health professionals that had previously completed the UK prescribing practice survey(7) and provided contact details were invited to take part in three case vignettes. Professionals were purposively selected based on their online survey responses to include a wide variation in anticoagulant prescribing in advanced CKD. Participants were selected to include a range different professionals including nephrology resident doctors, nephrology pharmacists and haematology resident doctors to understand variations in practice across professional groups, as highlighted in the survey.

There is no recommendation for the number of participants required but 15 participants were deemed to be sufficient to ensure that there would be data saturation(14). Twenty potential participants were e-mailed a study invitation and a copy of the participant information sheet. Anticipated time for participation was 30-60 minutes.

## Data collection

A Microsoft Teams or telephone meeting was arranged with the participants by the principal investigator (KP). Consent was audio recorded via an encrypted recording device prior to commencement of the vignettes. The case studies were then sent by e-mail so the participant could read the case and discuss how they would manage them. This was followed by questions to identify the reasons for their management plan. All discussions were audio recorded via an encrypted recording device as a separate file to the consent. Files were then transcribed verbatim.

## Data analysis

An inductive thematic analysis was undertaken as described by Braun and Clark(15). This analysis allows themes to be constructed by a six-phase framework. The framework includes familiarisation of the data by reading and re-reading the transcripts, generating initial codes, searching for themes, reviewing the themes to ensure that these fit the coded extracts and full dataset, defining and naming themes before finally writing the report(15). Both KP and PL familiarised themselves with the first 5 transcripts and developed a set of initial codes independently before discussing and refining them and applying them to further transcripts. Codes were then grouped by KP into themes and discussed with PL as part of a reflexive and iterative process. A meeting was held with JT, SM, PL and KP to review the themes and coding nomenclature and to agree the final themes. KP developed a table containing all themes with most relevant codes, a description of the theme and supporting quotes for review by all authors prior to writing the final paper. Reporting adhered to the COREQ checklist(16).

## Ethical approval

Ethical approval for this study was obtained from North West - Greater Manchester Central Research Ethics Committee REC reference 21/NW/0180, IRAS project ID: 281018

## Results

Of the 20 participants invited to take part, 15 accepted. This included six specialist renal pharmacists, six renal resident doctors and three haematology resident doctors. Only two of the haematology resident doctors had experience of working in a nephrology specific centre. Respondents were based across different regions of England with six from the North-West (5 NHS trusts), three from South-West (3 NHS trusts), three from the South-East (3 NHS trusts) and three from London (3 NHS trusts). Interviews were on average, 29 minutes in duration (16 – 43 mins).

There were five main themes that arose from the analysis, shown in Table 1. These over-arching themes were evidence, patient factors, knowledge and experience, team influence and systems and they were present across all the three vignettes. Findings are presented within these themes with quotations that exemplify these themes provided as examples.

**Table 1.**
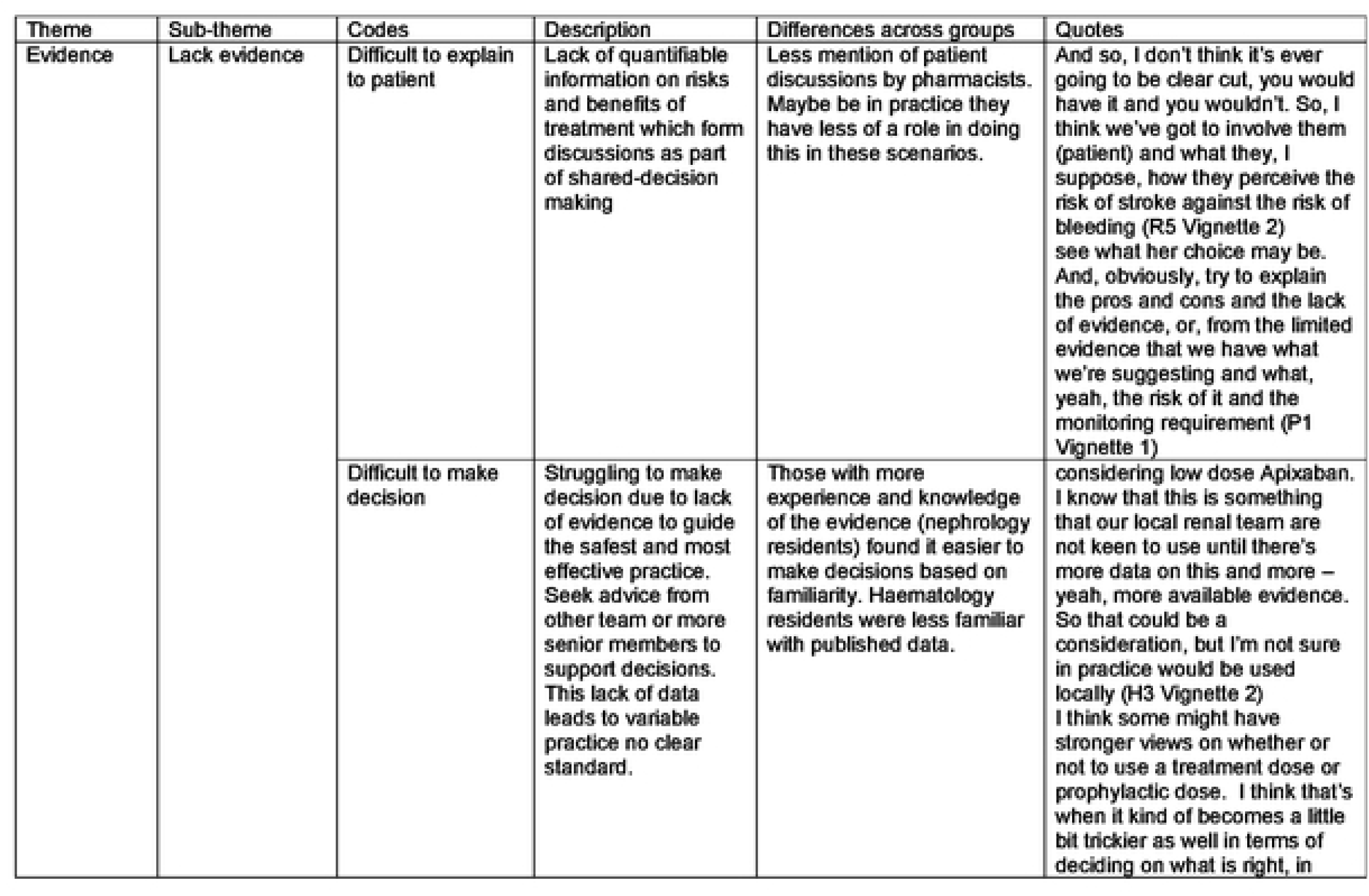
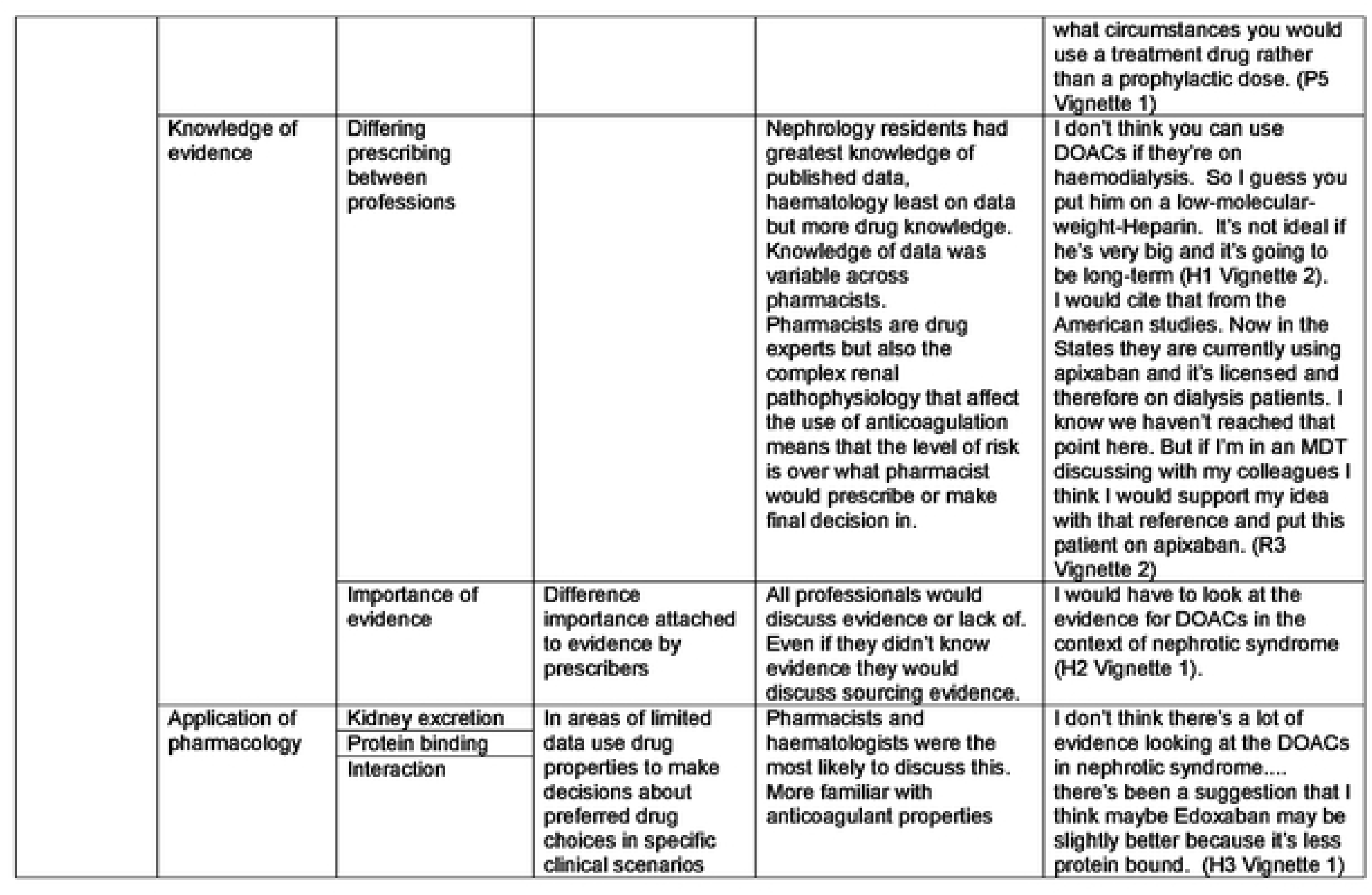
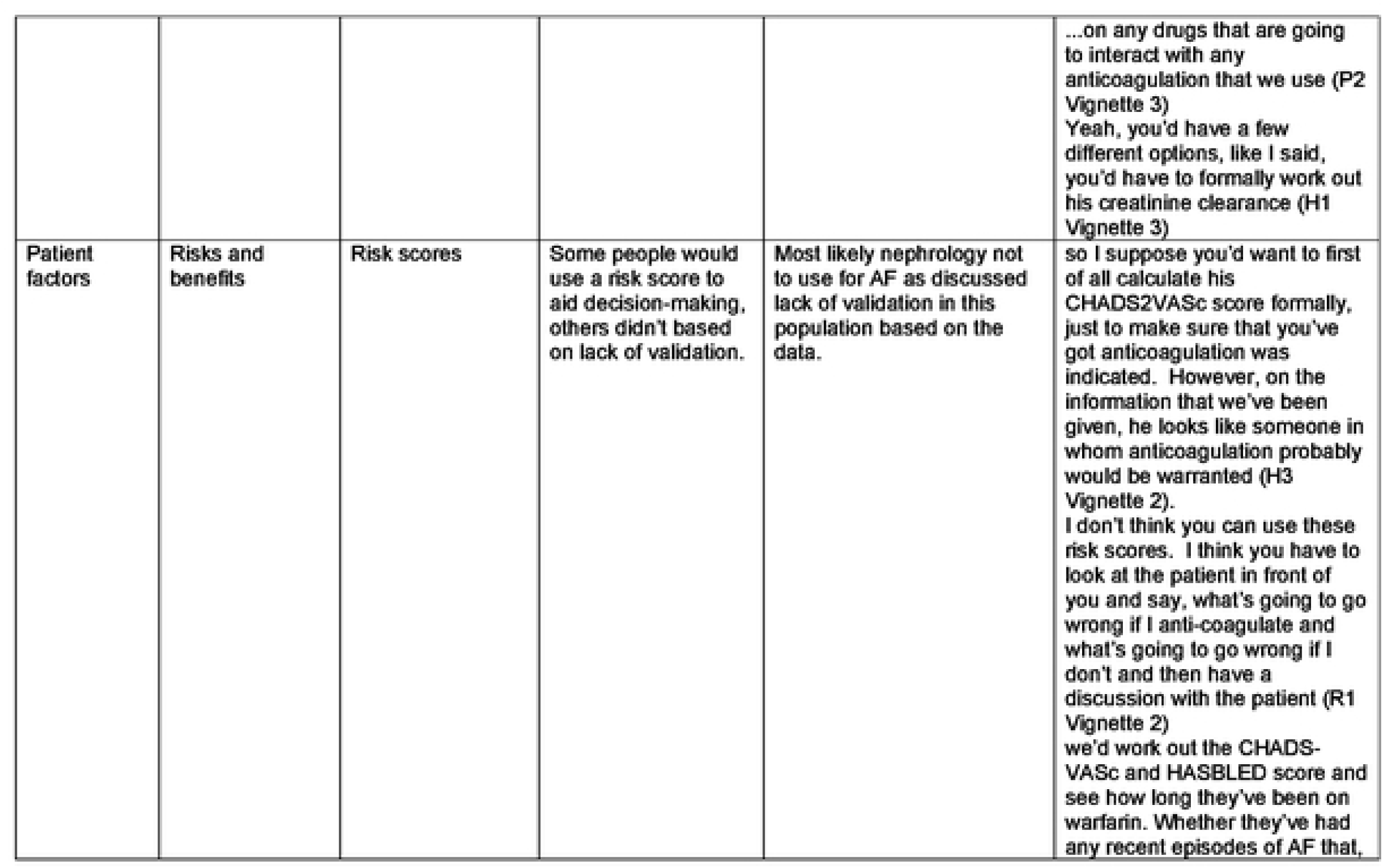
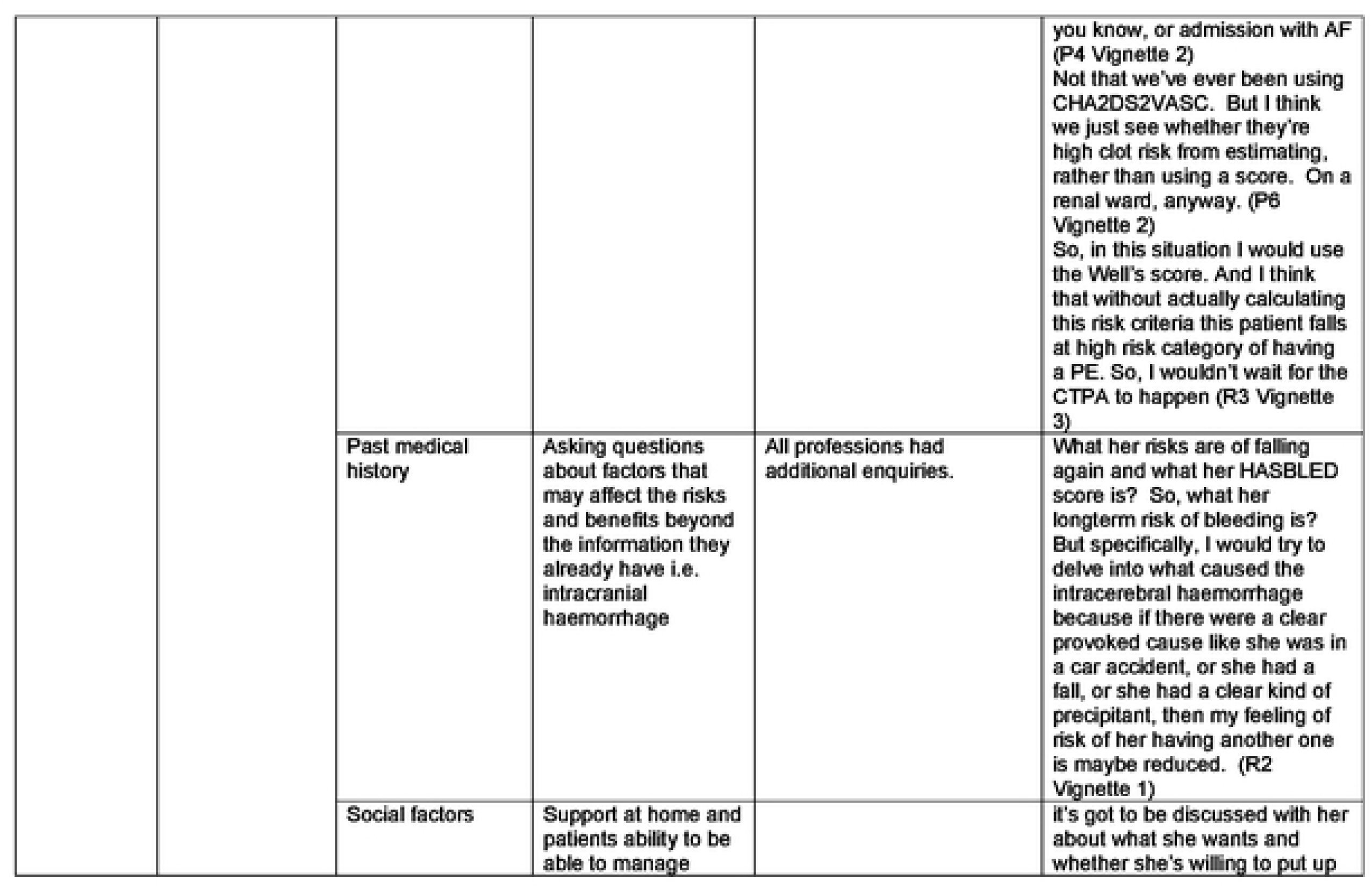
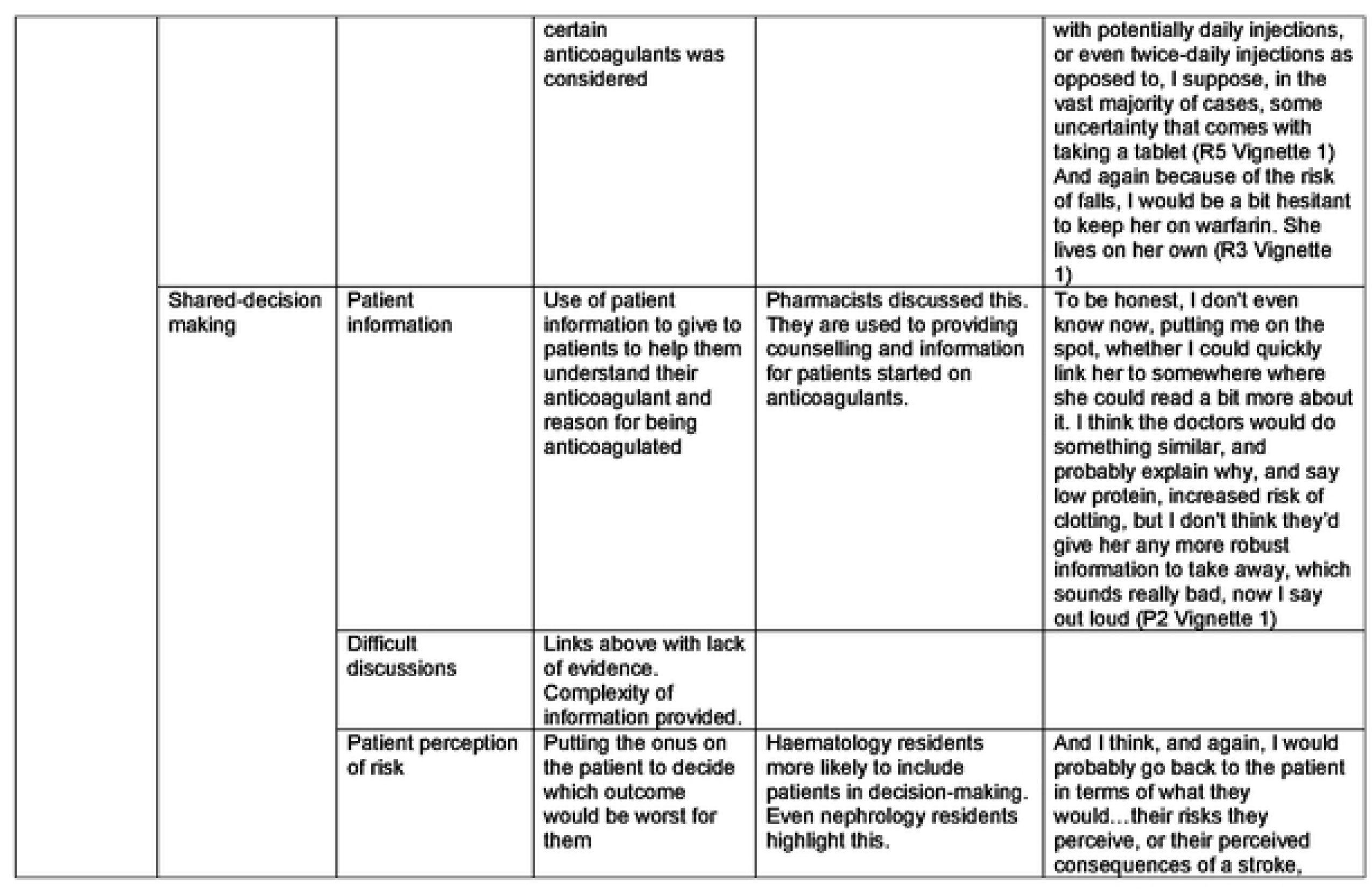
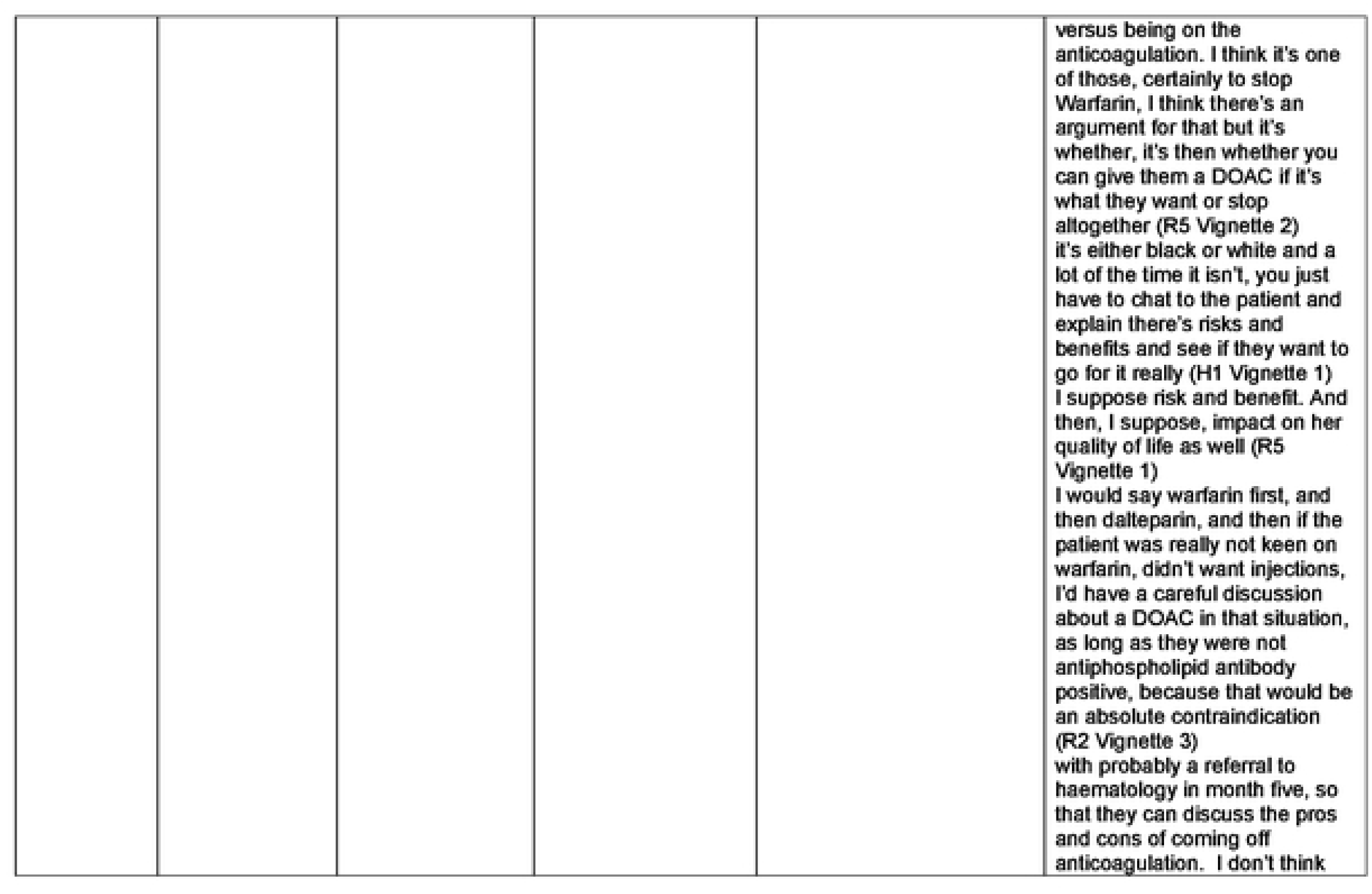
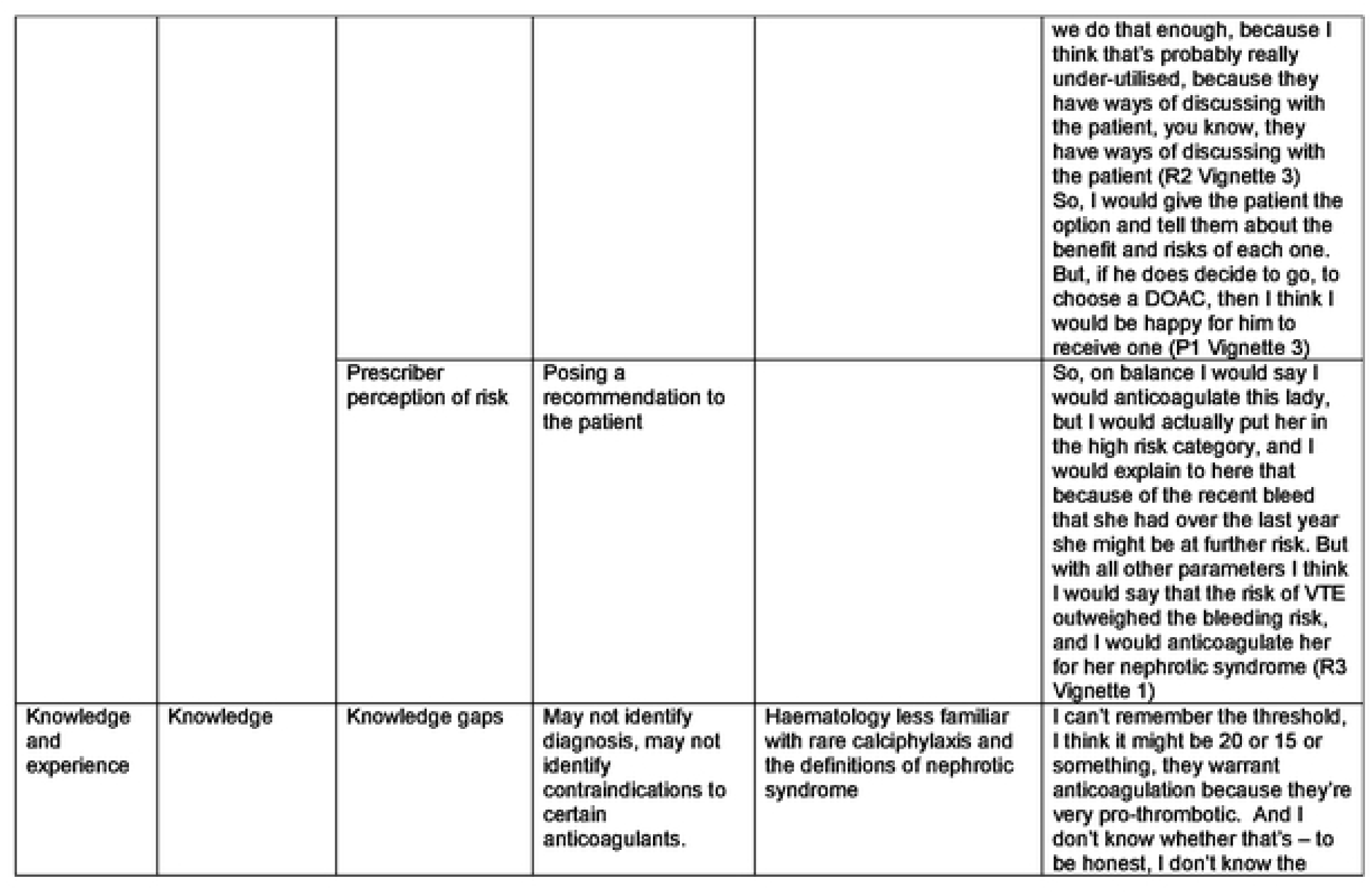
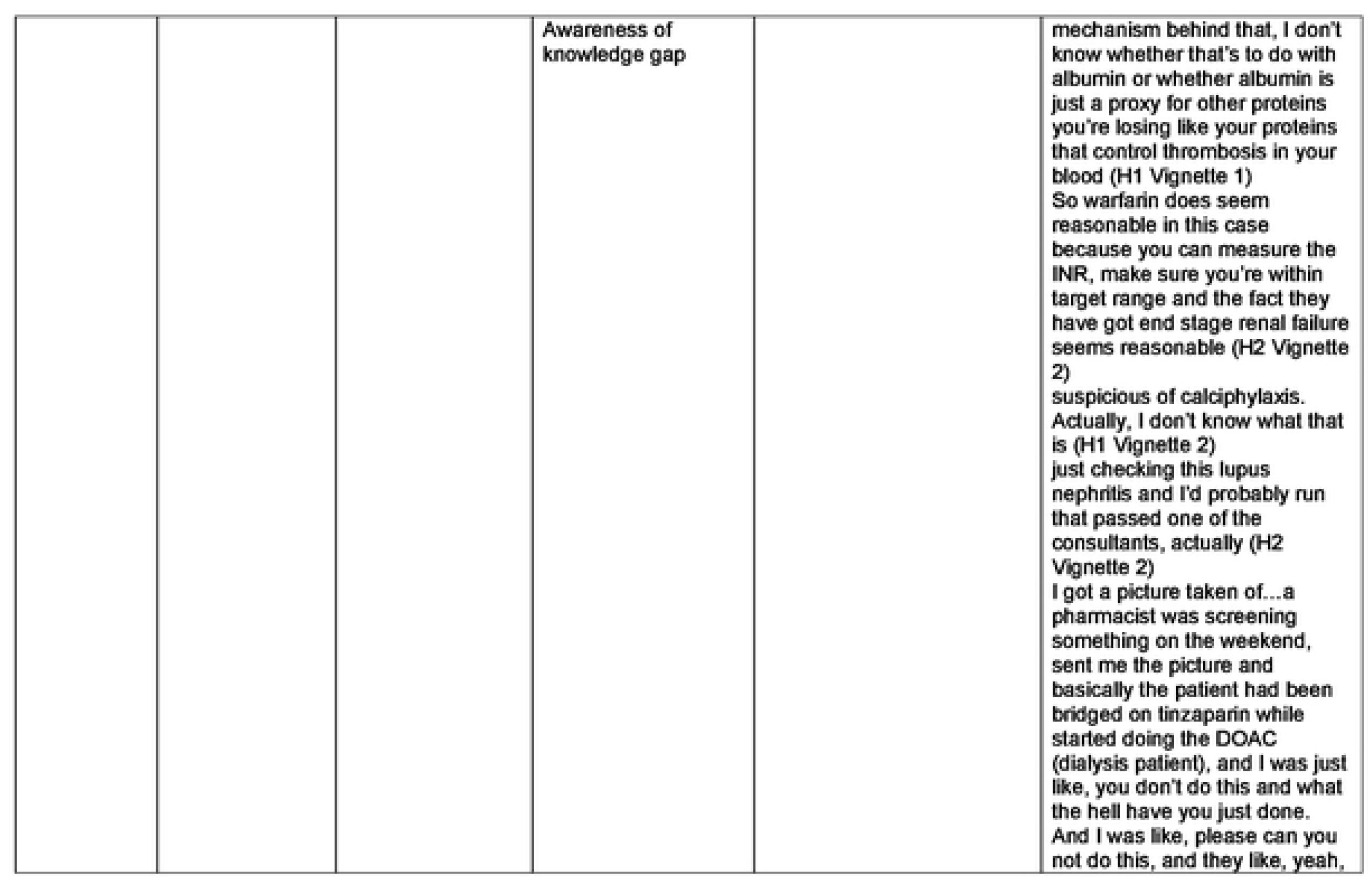
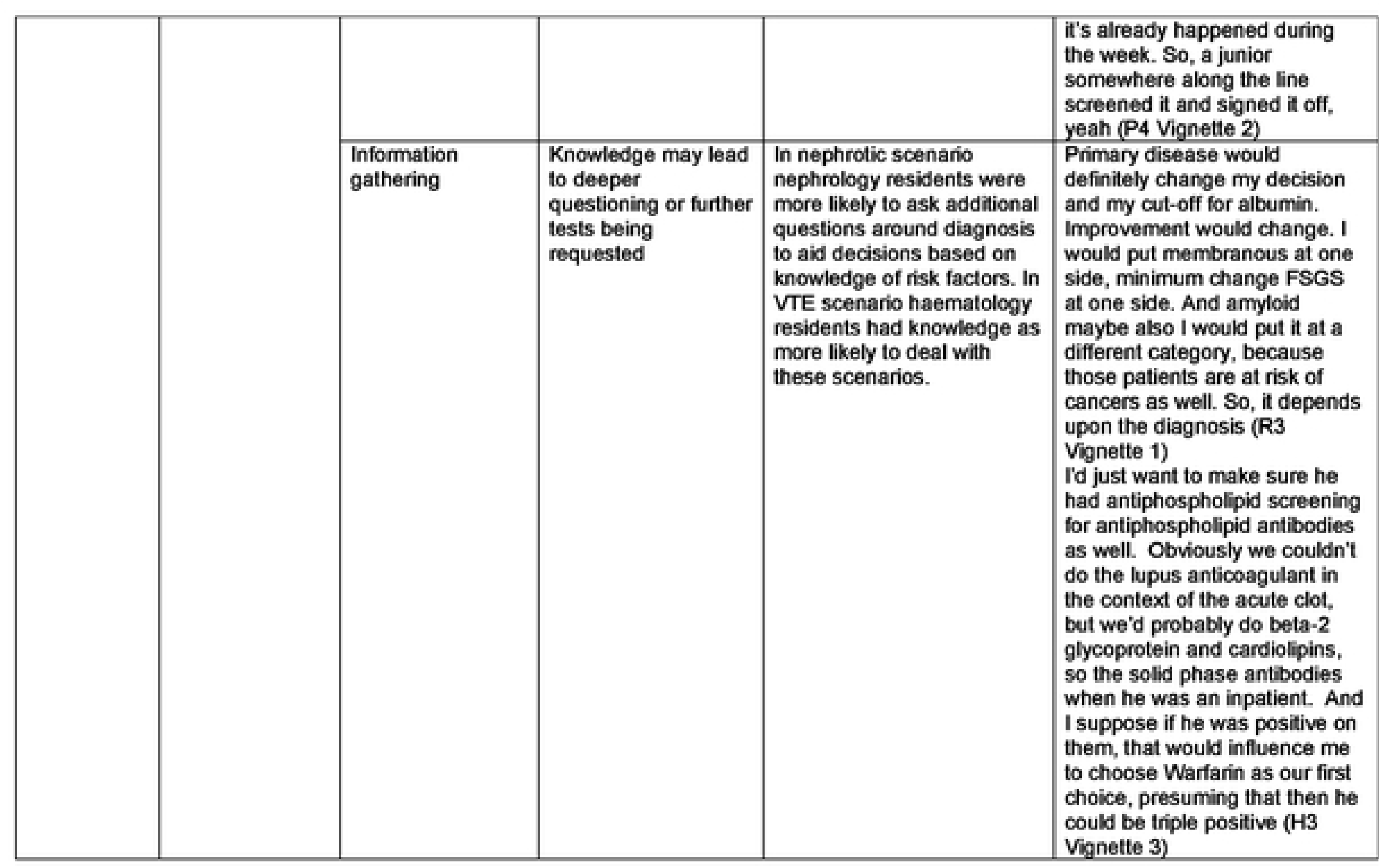
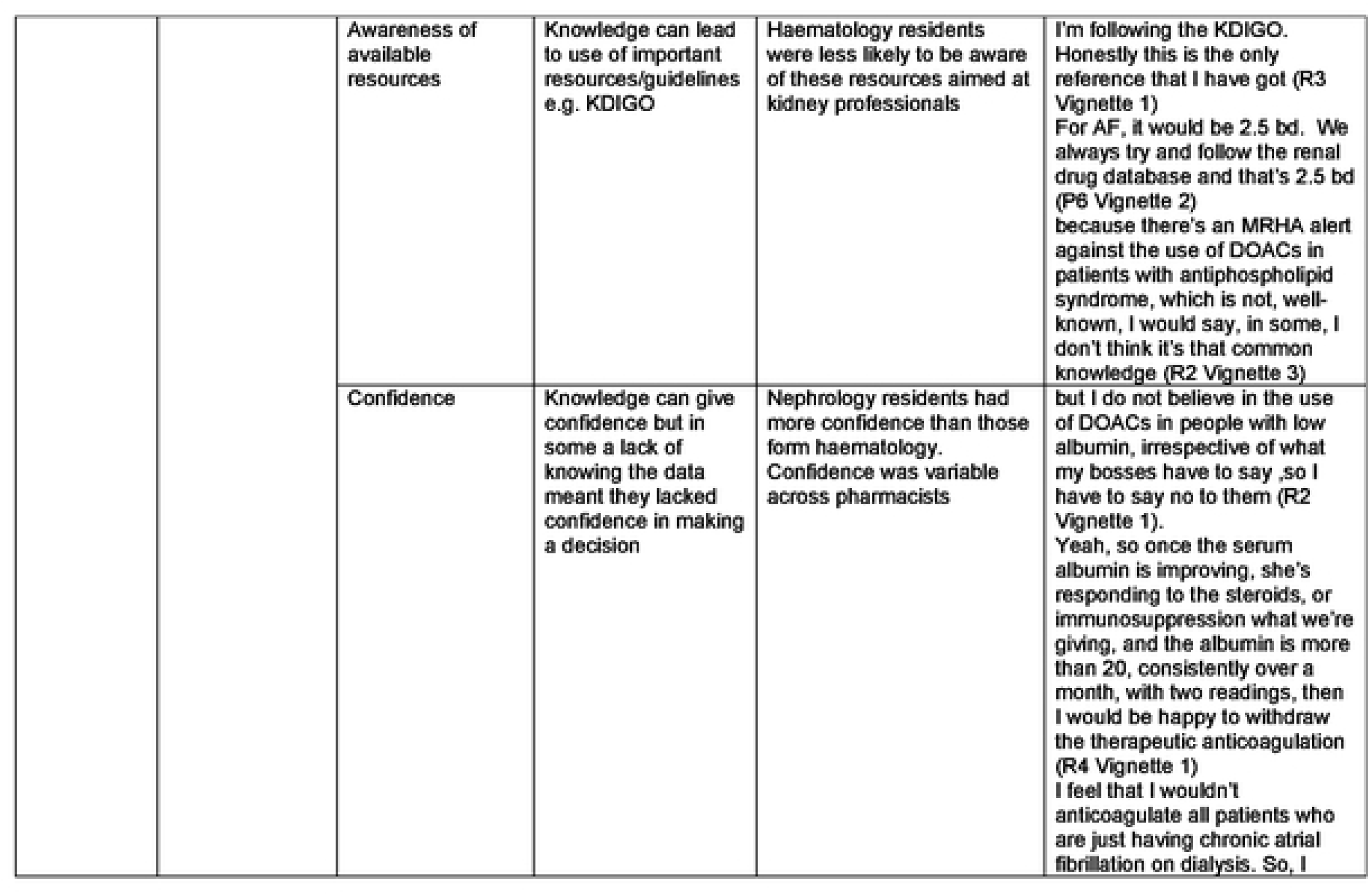
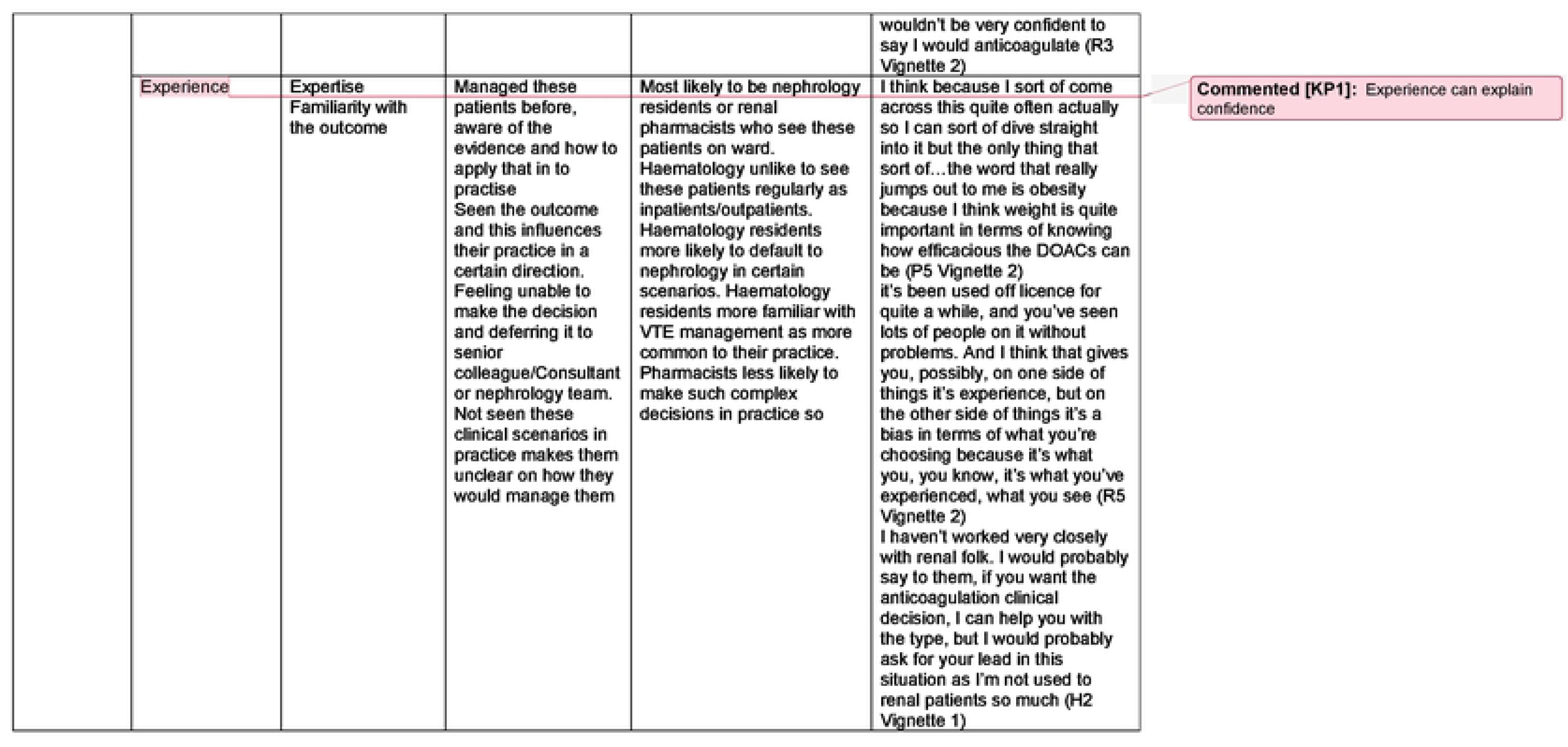
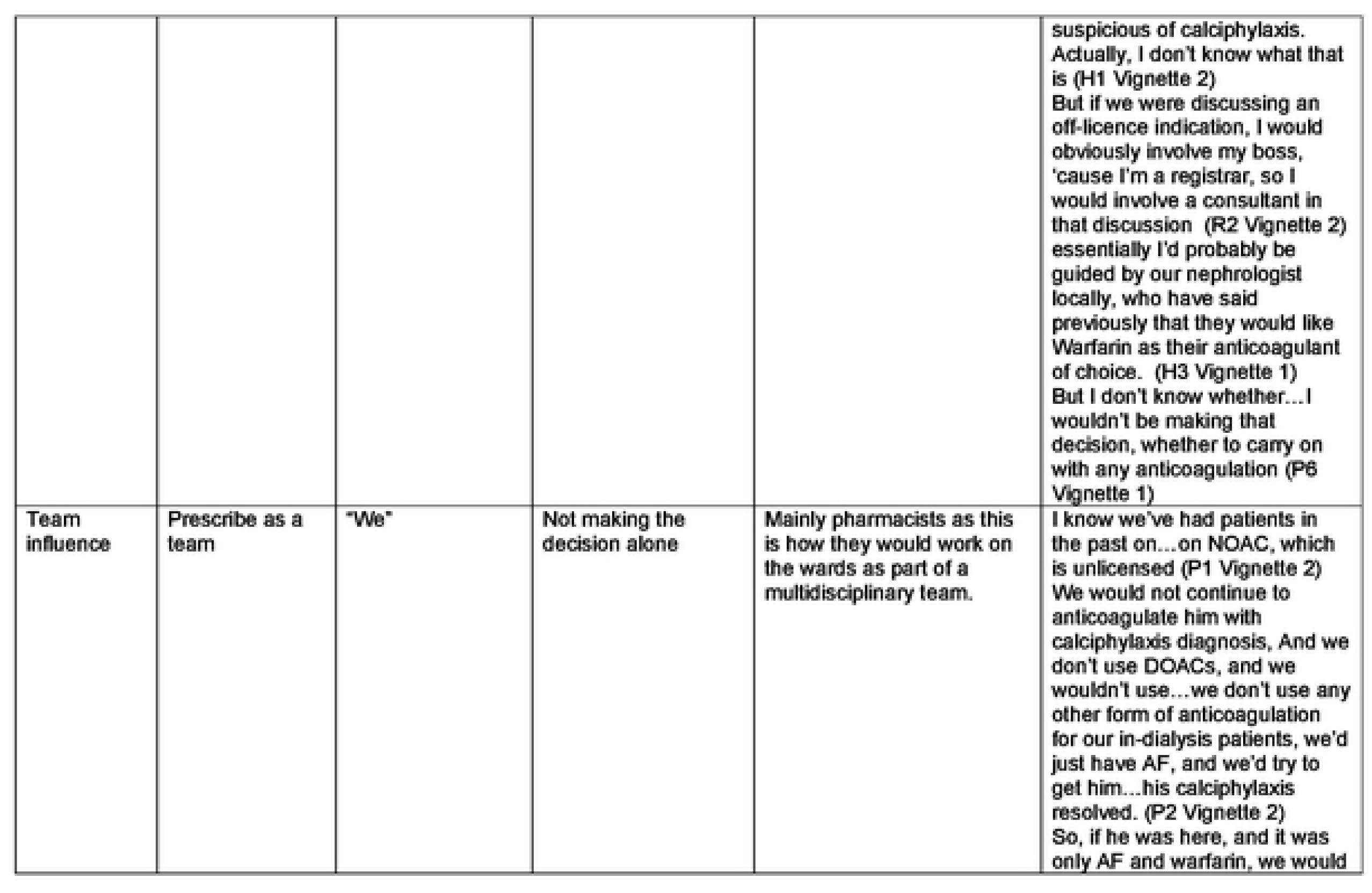
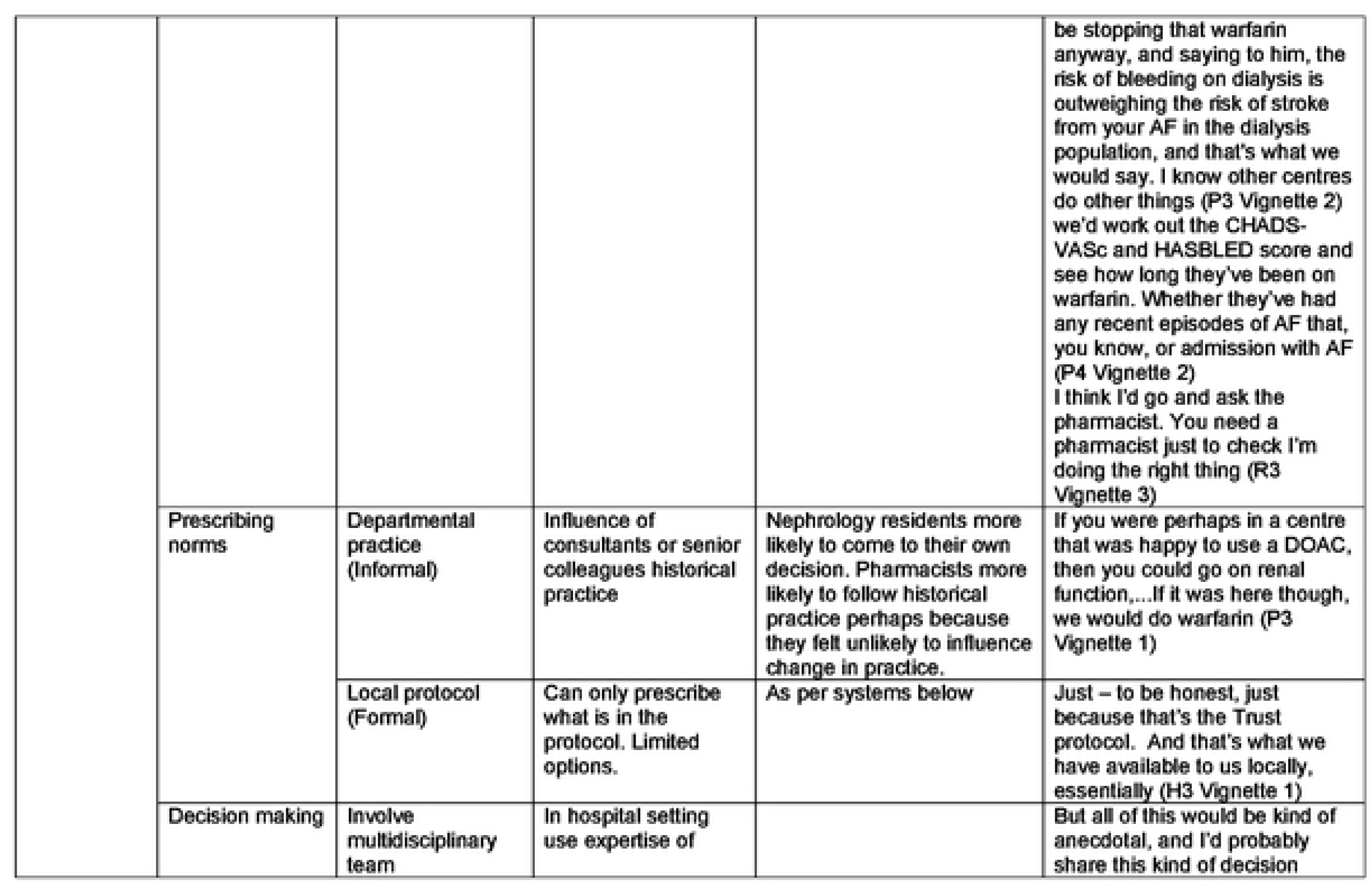
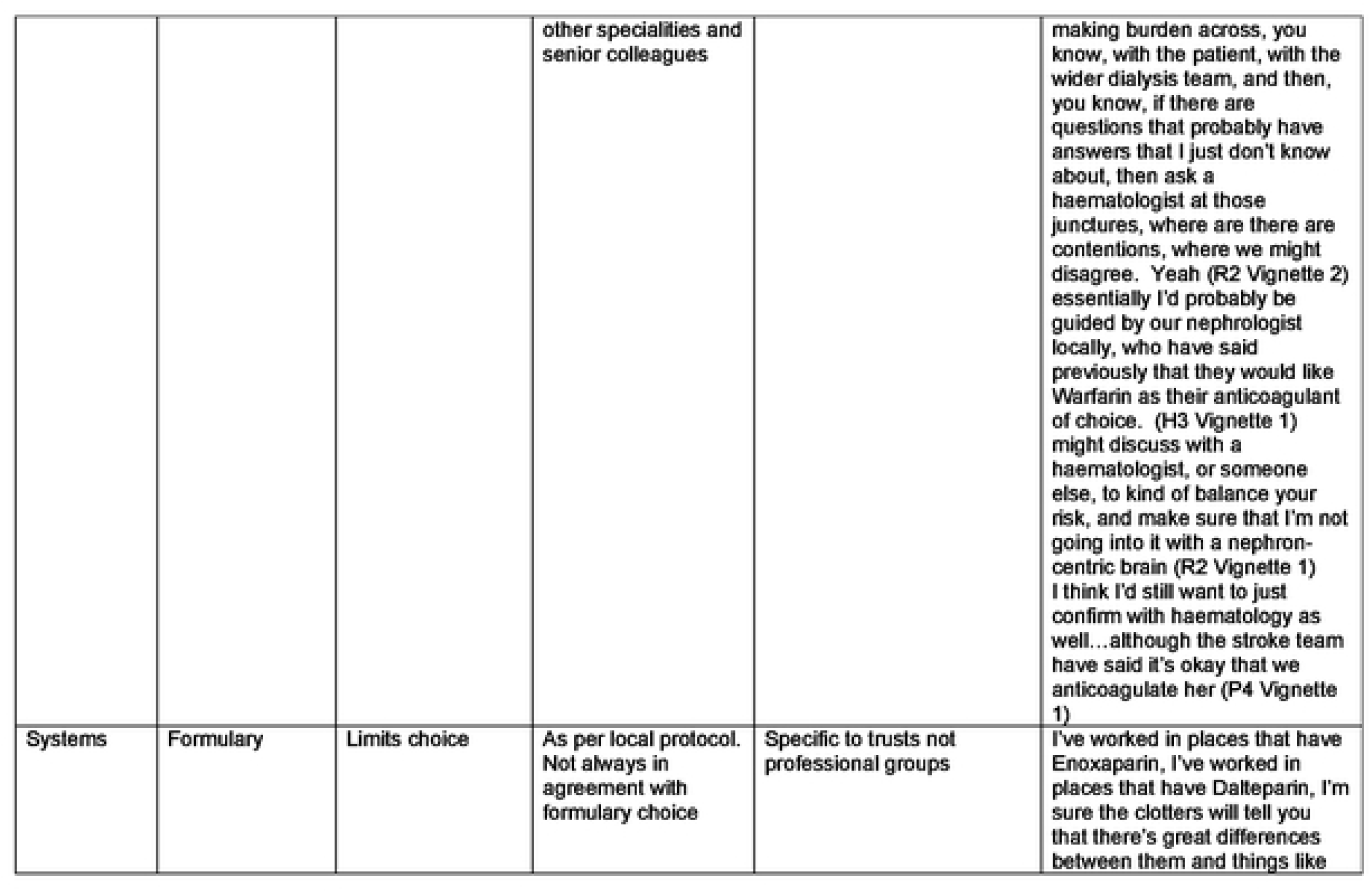
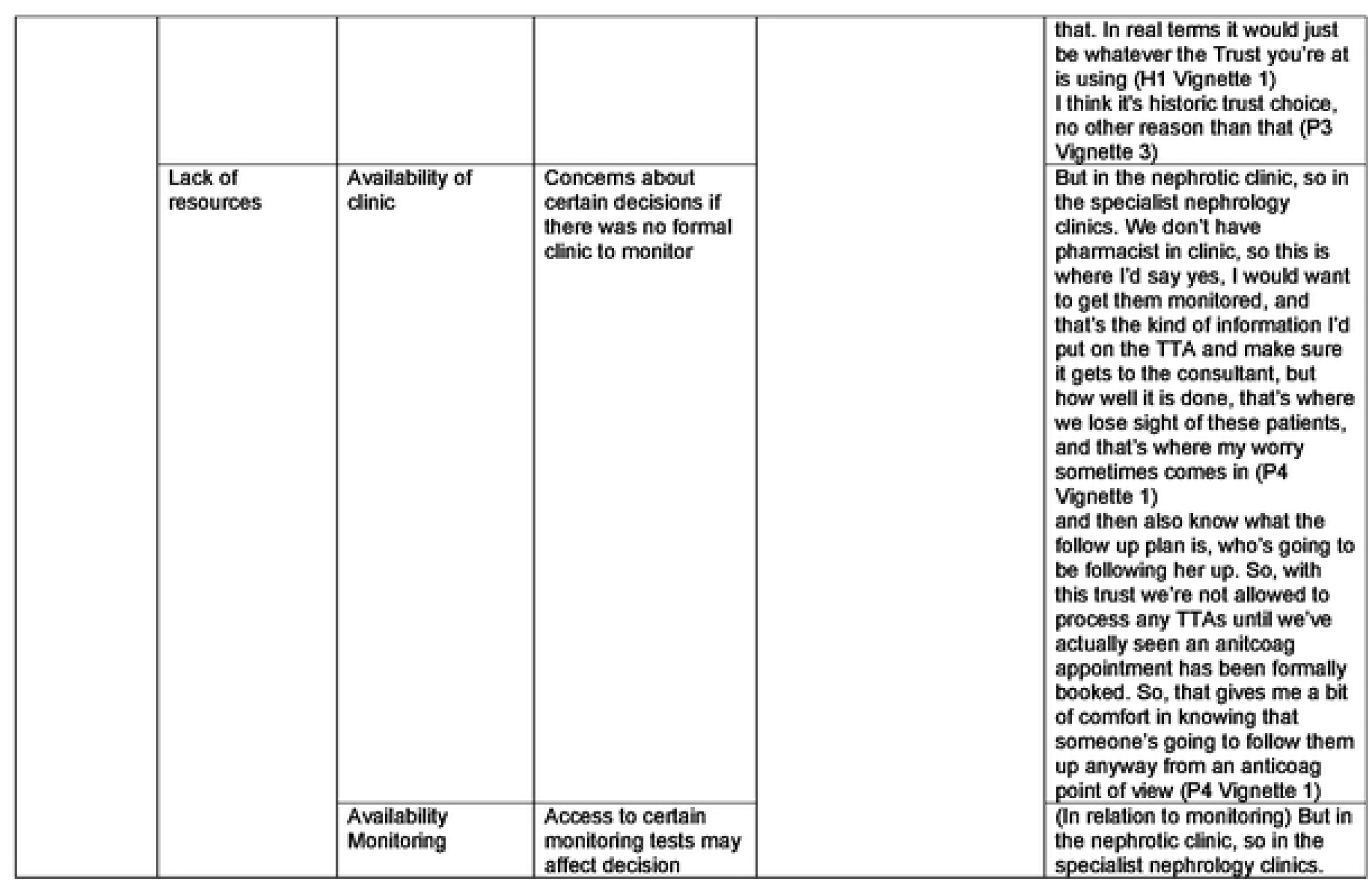
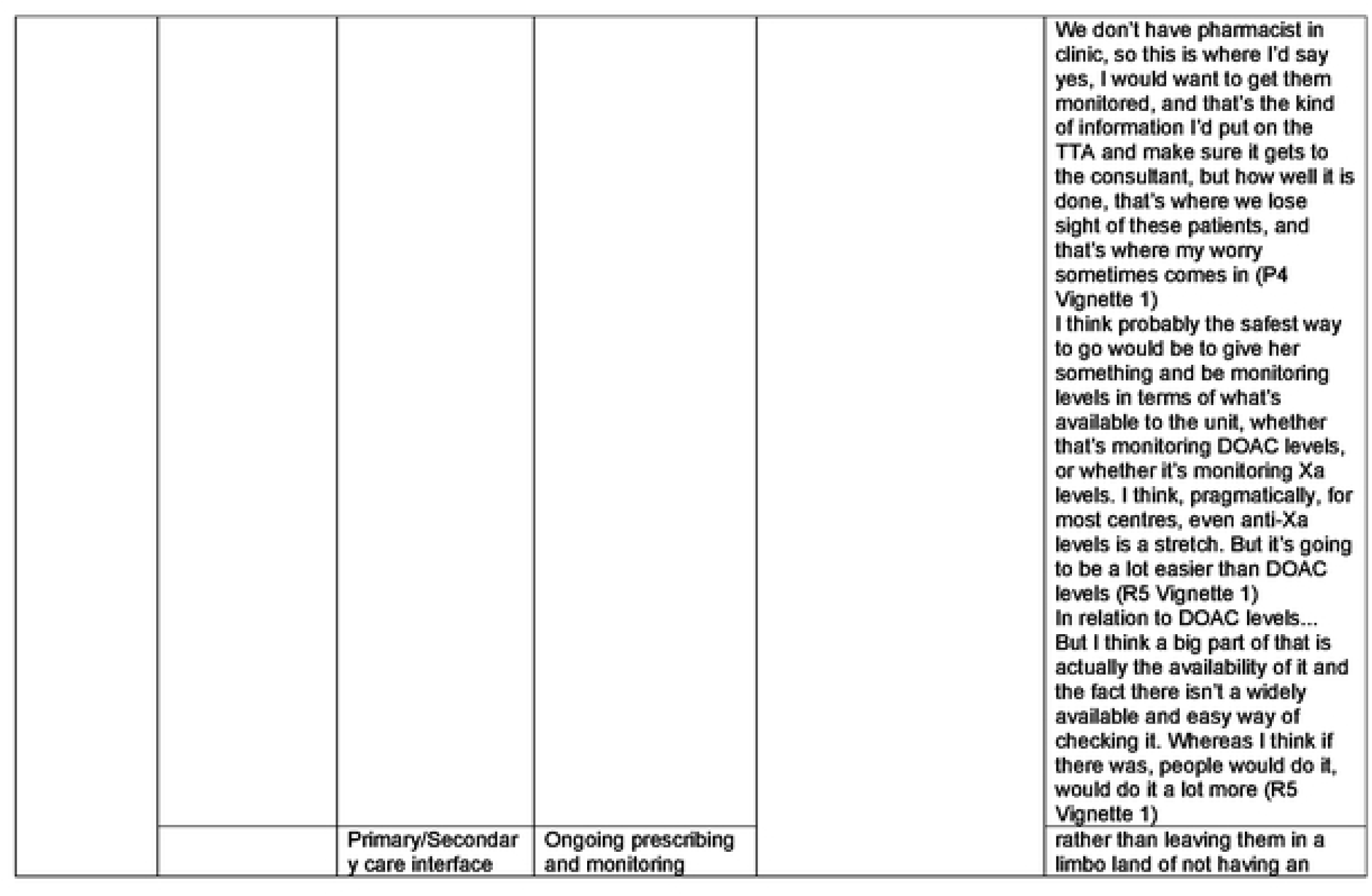
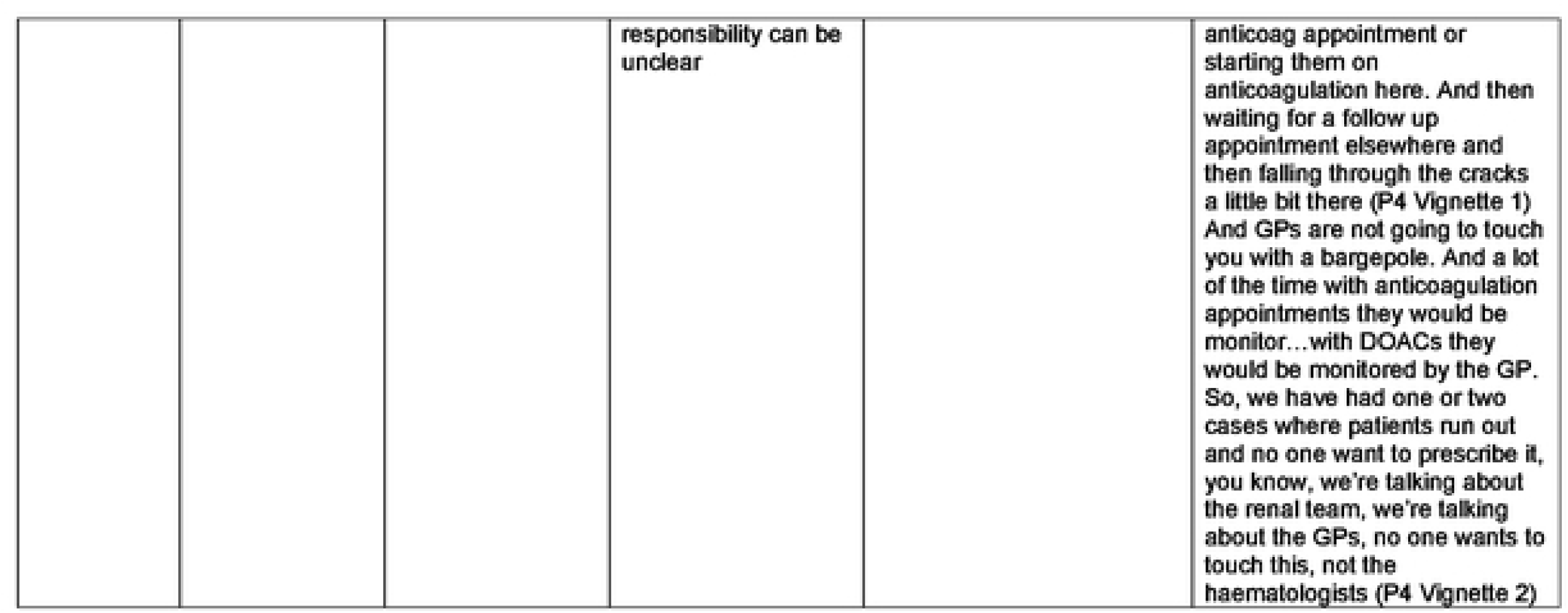
Themes relating to anticoagulant prescribing decisions in advanced kidney disease.

### 1. Evidence

As expected, all participants expressed difficulties in making decisions due a lack of available evidence as to the safest and most effective treatment within each scenario. This fundamental lack of evidence led to variable and uncertain prescribing practice within each of the vignettes, as there were no clear prescribing standards to adhere to. In response to this uncertainty, participants described how they would seek advice from the wider healthcare team or their senior colleagues to support their decision making

> *“I’d have to look up the guidance on dosing for it (apixaban) because I don’t think there is any recommended dosage. But I’d speak to haematology and see what they think. Also, a few of the dialysis consultants.” (R1 Vignette 2)*

Knowledge of the available evidence differed between professional groups, with nephrology residents and pharmacists having greatest knowledge of the limited data. However, all professions placed importance on the consideration of the available evidence when making prescribing decisions. In the absence of evidence, both haematology residents and pharmacists drew on their knowledge of the pharmacological properties of anticoagulant medication as a way of deciding upon the preferred medication choice. This approach was not seen in nephrology residents which was likely due to pharmacists and haematology residents’ expertise in anticoagulant pharmacokinetics.

> *“I don’t think there’s a lot of evidence looking at the DOACs in nephrotic syndrome. there’s been a suggestion that I think maybe edoxaban may be slightly better because it’s less protein bound.” (H3 Vignette 1)*

### 2. Patient factors

Assessing the risks and benefits of treatment for individual patients was important when making anticoagulant decisions and treatment choices. To assess the risks and benefits of anticoagulation in AF, haematology residents and nephrology pharmacists described using the risk assessment tools CHADS_2_-VA_2_Sc(17) for stroke and HAS-BLED(18) for bleeding risk – both widely used in clinical practice. However, nephrology resident doctors described the lack of validation of these risk tools in an advanced kidney disease population and instead based their decision on a holistic assessment of the patient and their co-morbidities rather than a risk assessment score.

The patient’s past medical history was a key consideration for all participants when assessing the risks and benefits of the prescribed medication. Participants often raised additional questions regarding the patient’s past medical history to determine current risks of treatment decisions:

> *“I would try to delve into what caused the intracerebral haemorrhage because if there were a clear provoked cause like she was in a car accident, or she had a fall, or she had a clear kind of precipitant, then my feeling of risk of her having another one is maybe reduced.” (R2 Vignette 1)*.

As part of a holistic approach to treatment risks, consideration was given to patients’ social circumstances, including their ability to manage anticoagulant treatment at home. This included, for example, the burden of multiple subcutaneous injections and concerns about the risks of adverse drug events if patients lived alone.

Shared-decision making featured heavily in the responses from all professional groups. However, discussing risks and benefits of anticoagulant treatment was highly problematic due to the lack of evidence of the risks and benefits of medication in an advanced-kidney disease population.

> *“I would probably go back to the patient in terms their risks they perceive, or their perceived consequences of a stroke, versus being on the anticoagulation. I think it’s one of those, certainly to stop Warfarin, but it’s whether, it’s then whether you can give them a DOAC if it’s what they want or stop altogether”. (R5 Vignette 2)*

All professionals highlighted the importance of elucidating the patients’ perceptions of risks of treatment. In these situations, decisions can be extremely challenging for patients who are effectively balancing the risk of stroke against the risk of bleeding from anticoagulation-both serious adverse events.

> *“I would probably go back to the patient in terms of what they would…their risks they perceive, or their perceived consequences of a stroke, versus being on the anticoagulation.” (R2 Vignette 2)*.

Pharmacists discussed the provision of patient information materials, including information about their condition, the rationale for anticoagulation as well as information about individual the anticoagulant. This was likely due to the role of pharmacists in the provision of anticoagulant counselling and information materials (e.g. yellow book for warfarin, DOAC booklets).

### 3. Knowledge and experience

Gaps in knowledge affected professionals’ decision making around anticoagulant use. In certain scenarios these knowledge gaps limited the understanding of a diagnosis which may have led to inappropriate recommendation of an anticoagulant. In Vignette 1, haematology residents did not recognise the criteria that defined nephrotic syndrome to aid decisions around initiating and stopping anticoagulants. In vignette 2, only some haematology residents had previously seen calciphylaxis, a rare condition associated with severe kidney disease, in a patient. However, several participants had an awareness of their knowledge gaps, and they described how in this scenario, they would refer to senior colleagues or undertake additional reading.

Knowledge led to more in-depth questioning to help further rationalise anticoagulant decisions. In vignette 1, nephrology residents wanted more information about the cause of nephrotic syndrome as it wasn’t provided until further down the question, whilst in vignette 3, haematology residents enquired about testing for antiphospholipid syndrome. Both of these are areas of familiarity for each speciality.

> *“Primary disease would definitely change my decision and my cut-off for albumin I would put membranous at one side, minimum change FSGS [focal segmental glomerular sclerosis] at one side. And amyloid maybe also I would put it at a different category, because those patients are at risk of cancers as well. So, it depends upon the diagnosis.” (R3 Vignette 1)*

Experience of managing similar anticoagulant scenarios led to development of expertise. By managing cases, respondents had learned what evidence was available and how it was applied in clinical practice to support anticoagulant decisions. Nephrology residents and pharmacists were more likely to see these patients in practice and had developed expertise. Haematology residents may not have come across these scenarios before, and their limited experience led to them seeking advice from the wider team when making decisions.

Professionals who had knowledge and experience of managing these clinical scenarios before had confidence in the management of anticoagulation as they had seen the outcomes of previous prescribing decisions:

> *“I think because I sort of come across this quite often actually so I can sort of dive straight into it but the only thing that sort of…the word that really jumps out to me is obesity because I think weight is quite important in terms of knowing how efficacious the DOACs can be.” (P5 Vignette 2)*

Nephrology professionals appeared to have greater confidence in making anticoagulant prescribing decisions than haematology residents and pharmacists. Confidence in making decisions varied across the pharmacist participants, likely because in clinical practice they are less likely to be making these decisions in isolation.

### 4. Team influence

Throughout the interviews, participants frequently used the term “we” when suggesting treatment decisions indicating that decision making was not an independent activity and would be undertaken as part of a multidisciplinary team. Pharmacists were the most likely to use this term and were unlikely to make the decision without wider multidisciplinary team input.

> *“We would not continue to anticoagulate him with calciphylaxis diagnosis, and we don’t use DOACs, and we wouldn’t use…we don’t use any other form of anticoagulation for our in-dialysis patients, we’d just have AF, and we’d try to get him…his calciphylaxis resolved.” (P2 Vignette 2)*

Working in a hospital setting meant that respondents would have access to a variety of specialists and they frequently described how they would include others in their anticoagulant decision making. For example, haematologists described how they would seek advice from nephrology and nephrology residents would seek advice from haematology. In the case of vignette 1, in which the patient had a history of intracranial haemorrhage, seeking advice from the stroke team on the safety of anticoagulation was highlighted. The use of pharmacists as a resource to review the anticoagulant prescription was also mentioned by nephrology resident doctors. Participants, particularly nephrology and haematology residents, described how they would likely to discuss complex decisions with their senior colleagues, such as their consultant. This approach shares the decision-making burden and broadens the range of considerations resulting in a more holistic response.

> *“[I] might discuss with a haematologist, or someone else, to kind of balance your risk, and make sure that I’m not going into it with a nephron-centric brain.” (R2 Vignette 1)*

Prescribing norms, which are established, generally accepted prescribing practices, were mentioned by respondents. These norms were based on prescribing practices that have been followed in their department for some time. Pharmacists were more likely to follow these practices as they felt that they would be unlikely to influence practice change, especially given gaps within the evidence.

> *“If you were perhaps in a centre that was happy to use a DOAC, then you could go on renal function,…If it was here though, we would do warfarin.” (P3 Vignette 1)*

### 5. Organisational Influences

The organisational context in which participants practised influenced anticoagulation decision making. These influences were usually beyond the participants’ control and included local trust formularies that restricted prescribing to specific anticoagulant agents. However, some participants disagreed with their local trust formulary believing that safer LMWH options existed for kidney patients, yet they felt unable to challenge this:

> *“Cause the contracting decision was made before they thought about the fact, we’re a kidney centre, and it’s something I’m not allowed to change. We can’t influence that.” (P2 Vignette 1)*

The availability of a specific outpatient clinic and associated follow up was raised as an important factor influencing participants’ decision making as the need for ongoing monitoring was deemed critical to ensure the ongoing safety of anticoagulant prescribing:

> *“We don’t have pharmacist in clinic [nephrotic clinic], so this is where I’d say I would want to get them monitored, and that’s the kind of information I’d put on the TTA [to take away discharge letter] and make sure it gets to the consultant, but how well it is done, that’s where we lose sight of these patients, and that’s where my worry sometimes comes in.” (P4 Vignette 1)*

Access to specific monitoring tests for certain anticoagulants was an influencing factor in some participants’ decision making as some centres did not have access to DOAC levels. Also, in some centres, anti-Xa for LMWH had to be approved by haematology before it could be undertaken which meant it was very difficult for them to undertake monitoring for therapeutic enoxaparin, resulting in safety concerns.

There was some discussion about the responsibilities for ongoing follow up to include prescribing and monitoring of anticoagulation and whether this would lie with primary or secondary care. In some instances, this left the patient in limbo as to where they were to obtain their next supply of medication:

> *“And GPs are not going to touch you with a bargepole. And a lot of the time with anticoagulation appointments they would be monitored…with DOACs they would be monitored by the GP. So, we have had one or two cases where patients run out and no one want to prescribe it, you know, we’re talking about the renal team, we’re talking about the GPs, no one wants to touch this, not the haematologists.” (P4 Vignette 2)*

## Discussion

This study has revealed the reasons for variability of anticoagulant prescribing decisions in advanced kidney disease and highlighted the main influences on these decisions: evidence, patient factors, experience and knowledge, team influence and systems. These are depicted in a diagram of influences in Figure 1. The study also highlighted how these influences differed by professional group.

**Fig 1.**
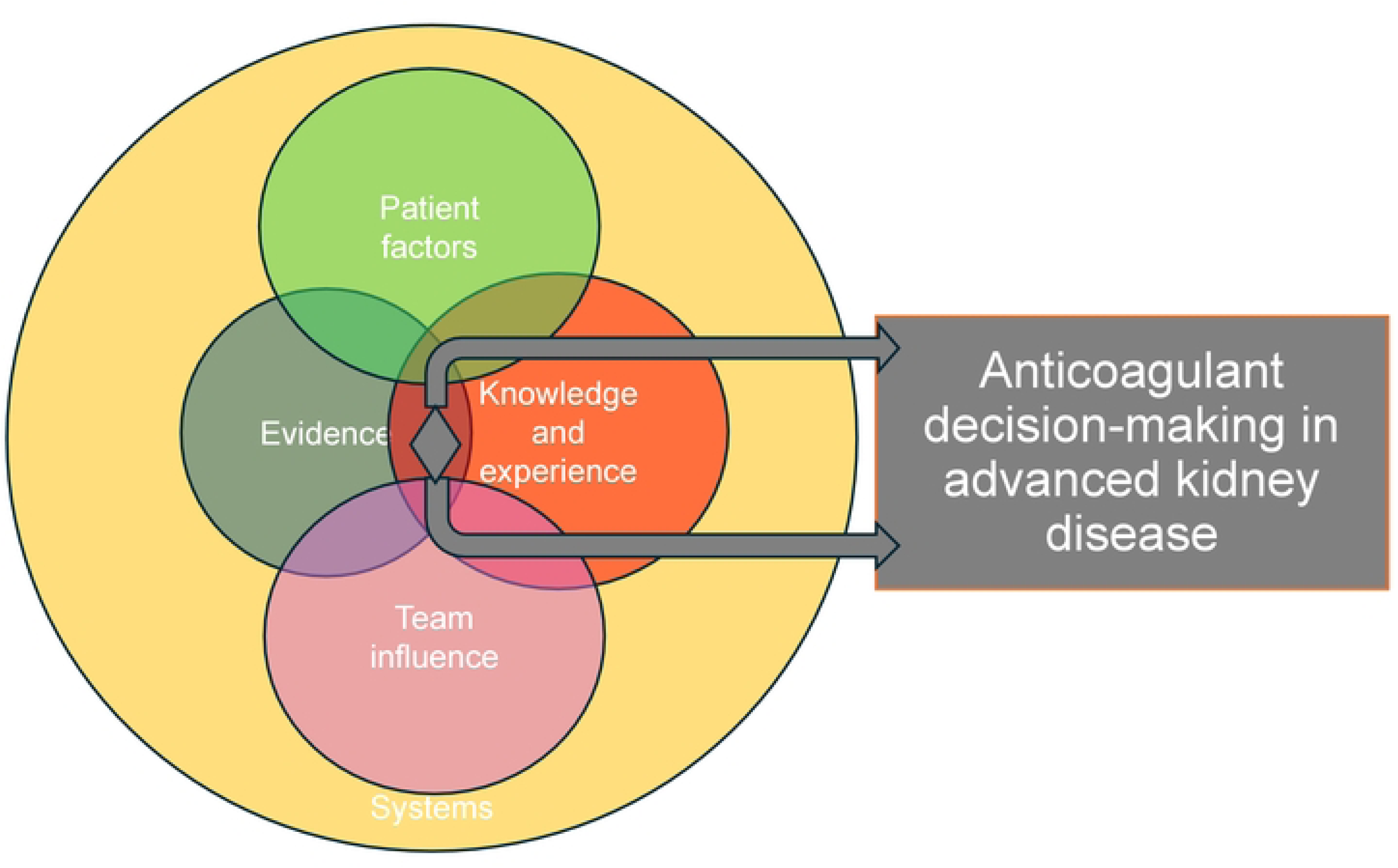
Influences diagram to show themes affecting decision-making

The absence of evidence or the poor-quality of evidence was highlighted by all participants which led them to rely on their previous experience or seeking advice from senior colleagues when making decisions. In an advanced kidney disease population, there are no RCTs for anticoagulant use in those with CrCl<30ml/min, leading to the availability of only a small number of studies subject to significant bias(4). To support prescribing practice going forward, studies examining anticoagulants in those with VTE and AF and CrCl<30ml/min and patients with nephrotic syndrome are required(4, 5). There is a need to develop research recommendations to support the use of anticoagulation in VTE and AF in advanced kidney disease and in nephrotic syndrome. This would include optimal drug dosing, therapeutic versus prophylactic anticoagulation in nephrotic syndrome, safety and efficacy of different anticoagulants in those with CrCl<30ml/min. There are ongoing studies investigating whether anticoagulation is appropriate in people with AF and CKD stage 5 or dialysis (NCT05679024). Unfortunately, a number of studies that had been set up have experienced problems with patient recruitment and have failed to provide evidence required to support practice. For example, VERDICT (NCT02664155) was a trial designed to explore reduced dose of DOACs in CKD stage 4 for treatment of VTE and did not recruit any participants, and the RENAL-AF study explored apixaban versus warfarin in dialysis, but recruitment was slow and funding was withdrawn (NCT03987711). Another approach may be the use of target-trial emulation, a research framework using observational data to emulate a hypothetical randomised controlled trial which is being used to overcome the costs and feasibility issues associated with real-world studies(19).

Patient involvement is key in all prescribing decisions(20) but, in the case of anticoagulant prescribing in advanced kidney disease, it is more difficult to involve patients in decision making due to the lack of evidence of treatment risks and benefits(10, 21). When assessing the risks and benefits of treatment, all prescribers gave consideration to patients’ comorbidities, social circumstances and current diagnosis. There was variability between professions as to whether they used risk scores when considering anticoagulation for stroke prevention in AF. Current evidence suggests none of the stroke and bleeding risks that are validated in a population with preserved kidney function are specific and sensitive enough to predict events in those with advanced kidney disease(22, 23) and nephrology resident doctors who were most likely to be aware of this data described these limitations. The National Institute for Health and Care Excellence (NICE) AF guidelines for the general population use a visual aid to represent the outcomes of bleeding and stroke to patients based on the risk score to support shared-decision making(21). This information is not available for people with advanced kidney disease, making it difficult for prescribers to discuss the risks and benefits of treatment with patients, hindering shared-decision making. Despite this, a previous study highlighted that people with advanced kidney disease wanted the risks of treatment to be presented clearly to them so they had awareness of what they may experience(10). Development of risk scores in this population could support shared-decision making with patients. Recently BLEED-HD, a bleeding risk score, was developed and validated for a dialysis population but showed low discrimination and was not validated in an AF population(24), perhaps the addition of other factors, such as biomarkers(25) or additional co-morbidities, may improve this score.

Within this study some of the renal resident doctors described how they would pose their recommendation to patients. This approach has been described in a previous study exploring patients’ perceptions of anticoagulation initiation in advanced kidney disease, where patients felt they were not involved in decisions and did not have a choice(10). Conversely, some patients prefer a non-participatory role in treatment decisions which leads to prescribers taking on sole responsibility(26). As part of shared-decision making there needs to be a discussion of what involvement the patient and family/carers would like in their treatment decisions as this will vary.

Maintaining knowledge is an important part of the prescribing process and is mentioned within the General Medical Council safe prescribing standards(27). The evidence for anticoagulation in advanced CKD is limited, poor-quality and often conflicting which makes it very difficult for professionals to keep on top of the literature. This was highlighted for all professionals, particularly haematology residents who may only have short training periods covering thrombosis and haemostasis. The majority of their training is in malignant haematology with limited opportunity or time to develop detailed knowledge of anticoagulant prescribing in patients with renal disease, particularly given the need to manage other complex areas of prescribing e.g. anticoagulation in liver failure, bleeding disorders. Development of a national guideline would support prescribing practice and summarise the current evidence base. Alongside this, development of a national training day on anticoagulation in advanced CKD for haematology registrars would support knowledge in this area. Nephrology professionals could also undertake such training via kidney professional organisations (e,g, UK Kidney association). Face to face training would be best to ensure participation in case discussions, but these could also be conducted online as a webinar and recorded for future reference. Haematology residents and pharmacists possessed good knowledge of drug pharmacokinetics which they described as part of their decision-making process. In nephrotic syndrome consideration to protein binding of drugs is important given that urinary protein losses may affect drug pharmacokinetics(28), and in those with CrCl<30ml/min the renal clearance of anticoagulants is important given LMWH and DOACs are renally excreted. Knowledge and experience develop over time which is why resident doctors from nephrology and haematology would often seek advice from their consultants. When decisions were made to use medicines off-label in a complex scenario, this would often result in participants seeking advice from the consultant as the responsibility would ultimately lie with them. Prescribers knowing their limitations and being aware of when to seek help and support is important to ensure that prescribing is safe(27) . Confidence comes after professionals have managed similar cases and gathered experiences of what the outcomes were, which will influence their future practice.

All professions discussed making decisions as part of and in conjunction with the multidisciplinary team (MDT). MDT working allows for a variety of expertise to be incorporated into the decision-making process and this is particularly valuable when there is uncertainty and complexity. MDT working has been shown to provide better outcomes for patients and provide more individualised care(29). However, what this study highlighted was that access to the multidisciplinary team was only available for inpatients and some participants felt that having additional support for outpatients would enhance the safety of anticoagulant use in people with kidney disease.

Organisational factors, beyond the control of individual prescribers, influenced prescribing decisions. Trust formularies restricted prescribing options to specific medications, particularly impacting on available LMWHs. The availability of a follow-up clinic and specific monitoring was a key system based influencing factor in anticoagulant prescribing decision making. Concerns were raised by participants about how safely an anticoagulant would be managed if there was not specific follow up via a designated clinic. The professional expected to follow up the patient may not have dedicated expertise to be able to manage these patients. To improve this, a clear plan for anticoagulation could be devised at the point of the initial review/discharge to ensure MDT input. This plan could then be followed by other team members, this plan could include specific monitoring required or parameters when anticoagulant should be discontinued.

Concerns were raised by prescribers regarding access to monitoring in certain NHS trusts. For example, anti-Xa was not always available for LMWH monitoring, or only available at the request of the haematology team. The Anti-Xa assay is readily available and is recommended by the manufacturers of LMWH in advanced kidney disease(30–32) so should be available to enhance safety in all NHS trusts. Some participants suggested DOAC level monitoring, which came up in all of the vignette scenarios. Its use is controversial because DOAC levels are not correlated with clinical outcomes and the level ranges are very large and therefore it is not recommended as a universal test (33).

Based on the findings of this study, Table 2 details future recommendations to support prescribing of anticoagulants in advanced kidney disease.

**Table 2.**
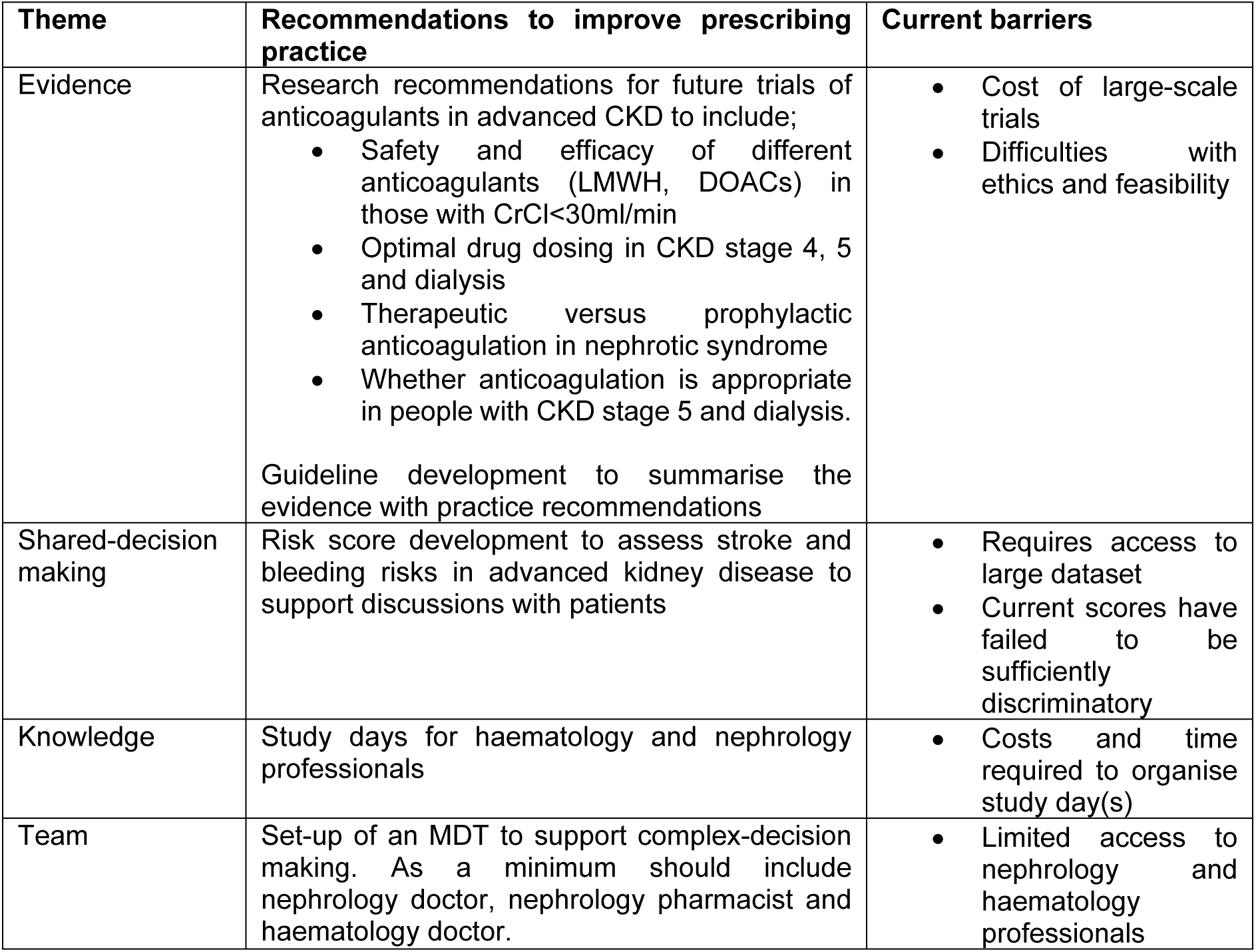
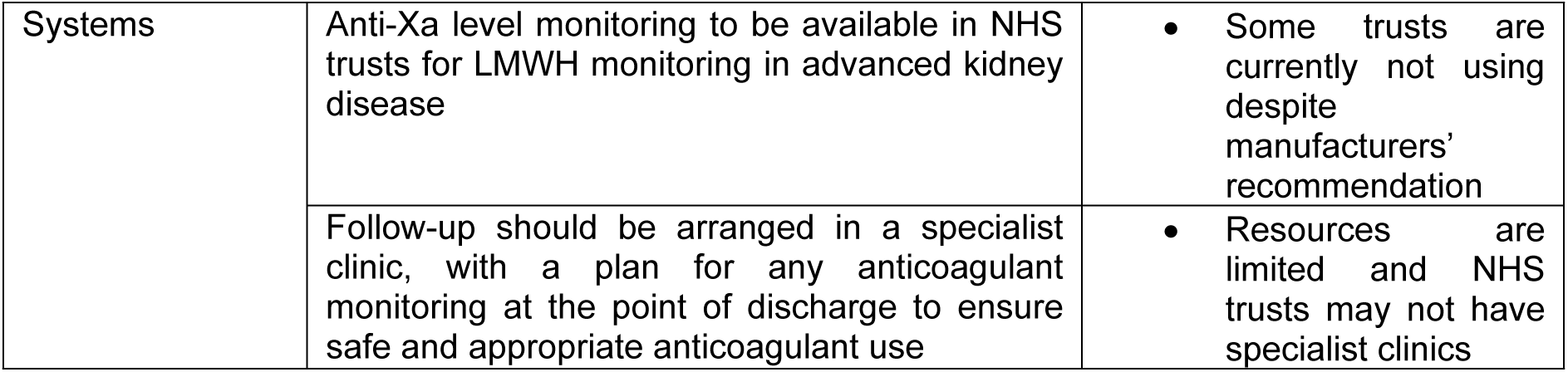
Recommendations to support prescribing practice of anticoagulants in advanced kidney disease.

## Strengths and weaknesses

This study purposefully recruited participants with variable practice from across multiple English NHS trusts, based on our previous survey of anticoagulant prescribing practice(7). Although the number of participants were small, data saturation was reached, with the same themes recurring across transcripts.

Vignettes are not real scenarios, so they do not allow for shared-decision making with patients or interaction with the wider healthcare team and therefore may not fully reflect the decisions that would be made in practice. However, they were effective at elucidating the key considerations and overarching influences on anticoagulant decision making by prescribers and the use of vignettes avoided the risk of generalisation and resolved the issue of recall when asking participants about their decision making.

## Conclusion

The prescribing of anticoagulation in advanced kidney disease is strongly influenced by the knowledge and expertise gleaned by prescribers’ previous experiences of anticoagulation scenarios and the incorporation of patient factors. Prescribers commonly seek opinion from senior staff and other members of the multiprofessional team to support their decision making. Decisions are also influenced by organisational factors that are beyond their individual control. Development of guidelines, delivery of education and good-quality evidence is needed to improve prescribing variation and ultimately the quality of care provided to patients with advanced kidney disease.

## Author Contributions

All authors were involved in conceptualisation of the study, design of study methodology and formal analysis of the data. KP lead on writing the draft manuscript with PL revising drafts. All authors reviewed and edited the manuscript. All authors agreed on the final manuscript.

## Data Availability

A summary of codes, including quotes, that led to data findings is available in Table 1. Due to ethical restrictions de-identified transcripts are not available unless there is a specific request to the corresponding author.

## Acknowledgments

**Kathrine Parker** was supported by the National Institute for Health Research (HEE/ NIHR ICA Programme Clinical Doctoral Research Fellowship, Miss Kathrine Parker, NIHR300545) to undertake a programme of research. The views expressed in this publication are those of the author(s) and not necessarily those of the NHS, the National Institute for Health Research or the Department of Health and Social Care.

